# ISO-101, a Wrist-Worn Isometric Counter-Manoeuvre Device, Improves Blood Pressure Recovery in Orthostatic Hypotension

**DOI:** 10.1101/2025.09.10.25335481

**Authors:** Lochlainn Connolly, Adam B. Smith, Neil Fawkes, Patrick I. McGowan, James Frith

## Abstract

ISO-101 is a novel wrist-worn device designed to deliver guided upper-limb isometric counter-manoeuvres via a retractable resistance tether with visual feedback. This proof-of-concept study evaluated feasibility, safety and preliminary efficacy in adults with orthostatic hypotension (OH). In a fixed-sequence, within-participant design, each participant completed three active-stand assessments: unassisted; device activation while supine immediately before standing; and device activation immediately after standing. The primary outcome was the proportion of responders, defined a priori as achieving, with device use versus the unassisted condition, ≥10 mmHg improvement in any one of resting blood pressure, nadir standing blood pressure at 30– 60 s, or reduction in the orthostatic blood-pressure drop (systolic or diastolic). The study was powered to test the responder proportion against a 20% benchmark derived from standard non-pharmacological care. Seventeen participants were enrolled (mean age 72 years; 77% male) and fourteen completed all assessments. Six of fourteen (43%) met the responder definition, exceeding the benchmark (exact binomial p=0.012). When the device was used immediately after standing, responders showed mean (SD) increases of +22.13 (16.29) mmHg systolic and +19.52 (12.47) mmHg diastolic compared with the unassisted stand; pre-stand activation yielded smaller, non-significant changes. No adverse events occurred. Usability was high, with 88% rating the device easy to use and 69% indicating they would use it in daily life. ISO-101 produced clinically meaningful improvements in orthostatic blood pressure in a substantial subset of patients, supporting progression to larger, controlled evaluations.

**WHAT IS ALREADY KNOWN:**

- Orthostatic hypotension (OH) is common and clinically significant, yet counter-manoeuvres are often poorly implemented in practice.
- Wearable assistance for standardised counter-manoeuvres has not previously been tested in OH.

**WHAT THIS STUDY ADDS:**

- In this proof-of-concept study, ISO-101, a guided wrist-worn device, achieved a clinically meaningful and statistically significant responder rate, exceeding that typically observed with standard care.
- Responders demonstrated marked increases in postural blood pressure, the device was rated as highly usable, and no adverse events were reported.

**HOW THIS STUDY MIGHT AFFECT RESEARCH, PRACTICE OR POLICY:**

- These findings highlight the potential for wearable-assisted therapy in OH and support the need for further clinical evaluation.

**TRIAL REGISTRATION NUMBER:** ClinicalTrials.gov: NCT06039410

## INTRODUCTION

Orthostatic hypotension (OH) is a chronic and frequently underdiagnosed condition characterised by a sustained reduction in systolic (≥20 mmHg) or diastolic (≥10 mmHg) blood pressure within three minutes of standing. Prevalence estimates vary by population and assessment criteria, but OH affects approximately 22.2% of community-dwelling older adults(1), with higher rates observed in those who are frail, institutionalised, or have neurodegenerative disease (2). Orthostatic Hypotension is associated with an increased risk of falls, cognitive decline, cardiovascular events, and all-cause mortality(3).

Current management options for OH include education, increased fluid and salt intake, physical counter-manoeuvres, and pharmacological agents such as fludrocortisone and midodrine(4). While non-pharmacological strategies are recommended as first-line interventions, real-world implementation is often limited by poor adherence, tolerability, and no intervention improving symptoms on standing(5).

Isometric physical counter-manoeuvres such as leg crossing, squatting, or upper limb contraction have been shown to transiently increase blood pressure by enhancing venous return and systemic vascular resistance (6, 7). However, these techniques rely on voluntary execution, which can be challenging for individuals with frailty, impaired coordination, or cognitive decline. There remains a need for accessible, standardised methods to support counter-manoeuvre-based management of OH in these vulnerable populations.

To address this unmet need, a novel wrist-worn medical device (ISO-101) has been developed to enable safe and effective delivery of isometric upper-limb contraction. The device incorporates a tethered resistance mechanism with embedded feedback indicators to guide users in performing the manoeuvre correctly and consistently during orthostatic stress.

This report presents preliminary findings from a single-arm proof-of-concept study evaluating the ISO-101 device in individuals with a clinical diagnosis of orthostatic hypotension. The study aimed to assess the feasibility, safety, acceptability, and short-term haemodynamic effects of the intervention.

## METHODS

This was a single-arm, prospective, proof-of-concept study designed to evaluate the safety, usability, and short-term physiological effects of a novel wrist-worn medical device (ISO-101) in individuals with orthostatic hypotension (OH). The study employed a fixed-sequence, within-subjects design, in which each participant completed three assessment periods involving postural blood pressure testing. The clinical investigation was conducted in accordance with ISO 14155:2020 and the principles of Good Clinical Practice. Approval was obtained from the Health Research Authority (IRAS ID: 1008987; REC reference: 23/YH/0202), and the study was prospectively registered on ClinicalTrials.gov (NCT06039410).

Participants were consecutive adults with OH from a single Falls and Syncope Service and Day Hospital in Northeast England. Eligibility for inclusion mandated they were aged 18 years or older and had a confirmed diagnosis of orthostatic hypotension, defined as a sustained drop in systolic blood pressure of at least 20 mmHg or diastolic blood pressure of at least 10 mmHg within three minutes of standing. Individuals were excluded if they had unstable arrhythmias, severe cognitive impairment, poorly controlled hypertension, or any musculoskeletal condition that would preclude safe use of the device. Written informed consent was obtained from all participants prior to any study procedures. Participants were asked to refrain from caffeine, nicotine and all medications, except for levodopa, before their appointment. They were instructed to have a light breakfast only. All participants provided written confirmation of informed consent. Orthostatic Hypotension was defined as neurogenic if the ratio of change in heart rate to change in BP upon standing was <0.49.

The investigational device, ISO-101, is a class 1 medical device developed to facilitate isometric contraction of the upper limb as a counter-manoeuvre to mitigate orthostatic blood pressure decline. It comprises a wrist-worn housing, connected to a retractable tether that provides resistance during contraction, alongside an integrated LED-based visual feedback system that guides the user in delivering appropriate effort over a specified duration. The force required is adjusted accordingly to the patients grip strength. The device is manually activated and designed for ease of use in older adults and those with physical limitations.

Each participant attended a single study visit during which they underwent three active stand assessments in a pre-determined sequence: (1) an unassisted active stand, performed without use of the device; (2) device activation while supine, immediately prior to standing; and (3) device activation immediately after standing. A minimum rest period of 10 minutes was observed between assessments to allow for cardiovascular stabilisation and to minimise sequence effects. All assessments were conducted in clinical research settings under continuous non-invasive haemodynamic monitoring using the Task Force Monitor (CNSystems).

The primary outcome was the proportion of participants classified as responders. A responder was defined a priori as achieving ≥10 mmHg systolic and/or diastolic BP improvement with device use relative to the unassisted active standing BP. This was assessed for improvement in any one of: (i) resting blood pressure, (ii) nadir standing blood pressure at 30–60 s, or (iii) reduction in the orthostatic blood-pressure drop. Criteria were applied to systolic or diastolic values. The primary objective was to determine whether the observed responder rate exceeded a prespecified threshold of 20%, a benchmark derived from reported efficacy rates of standard non-pharmacological interventions such as physical counter-manoeuvres(8).

Secondary outcomes included absolute changes in systolic and diastolic blood pressure across the three assessment periods, time to blood pressure recovery, and heart rate response. In addition, participants completed a structured questionnaire to evaluate the acceptability and usability of the device following the final assessment. Any adverse events or device-related complications occurring during or immediately after device use were documented in accordance with clinical safety procedures.

Statistical analysis focused on descriptive and exploratory inferential methods. The responder rate was evaluated using an exact binomial test to assess whether it exceeded the 20% threshold under the null hypothesis. Paired comparisons of haemodynamic outcomes between assessment conditions were conducted using parametric or non-parametric tests, depending on data distribution. All analyses were conducted on a complete-case basis and interpreted within the context of a proof-of-concept study.

## RESULTS

Seventeen patients were enrolled in the study, comprising 13 males (77%) and 4 females (23%). The mean age was 72 years (SD 7.8, range 51 to 84 years), with a mean body mass index of 23.4 (SD 2.99) kg/m^2^. Baseline supine haemodynamic parameters were a mean systolic blood pressure of 119.2 (SD 14.95) mmHg, mean diastolic blood pressure of 80.9 (SD 15.0) mmHg, and mean heart rate of 76.7 (SD 22.3) bpm. Twelve participants (86%) had neurogenic orthostatic hypotension, two patients with standard OH (24%). Concomitant medication use included levodopa in 6 patients (35%), fludrocortisone in 4 (24%), beta-blockers in 3 (18%), and midodrine in 2 (12%).

Three patients (18%) were excluded from the analysis due to incomplete or unusable haemodynamic recordings affected by movement artefact during device use, leaving 14 patients for evaluation of the primary endpoint. Six of these 14 participants (43%) met the predefined responder criteria, which exceeded the prespecified clinical meaningful benchmark of 20% based on reported standard-of-care performance. The exact binomial test indicated this difference was statistically significant (p = 0.012) of the 14 participants. Among responders, three showed improvements in both systolic and diastolic blood pressure, two in systolic only, and one in diastolic only.

Responder status was associated with significantly greater improvements in blood pressure compared with non-responders (see Supplementary Figure 1). For systolic blood pressure, the mean change between the unassisted stand and device use immediately after standing was +22.13 (SD 16.29) mmHg in responders versus in non-responders (p = 0.0012). For diastolic blood pressure, the corresponding difference was +19.52 (SD 12.47) mmHg (p = 0.0015).

Device use while supine prior to standing produced a mean systolic increase of 18.58 (SD 7.05) mmHg and a diastolic increase of 14.02 (SD 11.51) mmHg among responders however, statistical significance was not demonstrated.

No adverse events or device-related complications were reported. Patient-reported experience was favourable, with 76% of participants scoring overall satisfaction greater than four on the 7-point scale. Most participants found the device easy to use (88%) and useful (69%) and expressed useful to use it in daily life.

## DISCUSSION

This proof-of-concept study provides preliminary evidence that a novel wrist-worn device (ISO-101, ISOTECH Ltd) can improve orthostatic blood pressure responses in a subset of patients with clinically diagnosed orthostatic hypotension. The proportion of participants meeting the predefined responder criterion was 43%, which exceeds the benchmark responder rate of 20% reported for standard non-pharmacological counter-manoeuvres in the literature(8). Importantly, these improvements were observed in a cohort with a high prevalence of neurogenic orthostatic hypotension, a group often refractory to lifestyle and pharmacological measures.

Device use immediately after standing produced the largest improvements in both systolic and diastolic pressures among responders, with mean increases of 22 mmHg and 19 mmHg, respectively, much greater than the primary endpoint benchmark of >10mmHg (5). In contrast, use while supine prior to standing produced more modest changes, suggesting that active engagement during the period of orthostatic challenge is critical for optimising benefit. These differences are consistent with physiological effects described for voluntary isometric counter-manoeuvres, which operate by increasing systemic vascular resistance and venous return(7).

The observed responder rate and the magnitude of blood pressure improvement are notable given the advanced age, comorbidity burden, and concomitant medication use in the study population. Previous reports indicate that adherence to traditional counter-manoeuvres is poor in such populations, partly due to impaired mobility, coordination, or cognitive function (9). ISO-101 offers a standardised and accessible means of delivering a counter-manoeuvre, with integrated feedback to guide correct technique, potentially addressing these barriers. The high usability scores and absence of adverse events further support its feasibility in routine use.

Several limitations warrant consideration. The study employed a small sample size and a non-randomised sequential design, which may introduce order effects and limit generalisability. The absence of long-term follow-up precludes conclusions regarding sustained efficacy or adherence. In addition, the fixed responder threshold, while based on literature and clinical rationale, may not capture the full spectrum of meaningful patient benefit. Continuous beat-to-beat monitoring was used to standardise physiological assessment, but real-world use is likely to be more variable.

Future research should aim to validate these findings in larger, randomised controlled trials, including patient-centred outcomes such as fall frequency, symptom burden, and quality of life. Comparisons with established counter-manoeuvre techniques and pharmacological agents would further clarify the role of ISO-101 within the therapeutic pathway for orthostatic hypotension. Investigation into optimal timing, contraction intensity, and integration with other interventions may also enhance device performance.

In summary, this study demonstrates that ISO-101 can safely and effectively improve orthostatic blood pressure recovery in a proportion of patients with orthostatic hypotension, achieving a responder rate exceeding that expected from standard guidance. These findings support further evaluation of the device as a practical and scalable adjunct to current management strategies.

## CONCLUSION

In this proof-of-concept study, use of the ISO-101 device was associated with clinically meaningful improvements in orthostatic blood pressure responses in a proportion of patients exceeding the expected benefit from standard non-pharmacological interventions. The device was well tolerated, easy to use, and demonstrated no safety concerns. These preliminary findings support the potential role of ISO-101 as an accessible adjunct to existing management strategies for orthostatic hypotension and justify further evaluation in larger, randomised controlled trials with longer-term follow-up.

## Data Availability

All data produced in the present work are contained in the manuscript

## DECLARATIONS

### ETHICAL APPROVAL

The study received ethical approval from the Health Research Authority and Yorkshire & the Humber – Bradford Leeds Research Ethics Committee (REC 23/YH/0202; IRAS ID 1008987). All participants provided written informed consent. The investigation was conducted in accordance with ISO 14155:2020, ICH-GCP and the Declaration of Helsinki, and was prospectively registered on ClinicalTrials.gov (NCT06039410).

### AUTHOR CONTRIBUTIONS

J Frith (Principal Investigator) and the Newcastle team conceptualised the study, designed the protocol, and conducted the clinical investigation independently. A Smith served as an independent statistician and performed the statistical analysis. The Isotech team (N Fawkes, L Connolly, I McGowan) supported the study with device expertise, training, and technical input. All authors contributed to data interpretation, critically revised the manuscript for intellectual content, and approved the final version.

## ACKNOWLEDGEMENTS

We thank the participants and the clinical staff within the Falls and Syncope Service, Newcastle upon Tyne Hospitals NHS Foundation Trust, for their support and assistance. We also acknowledge Innovate UK for funding support.

## FIGURES AND TABLES

**TABLE 1.**
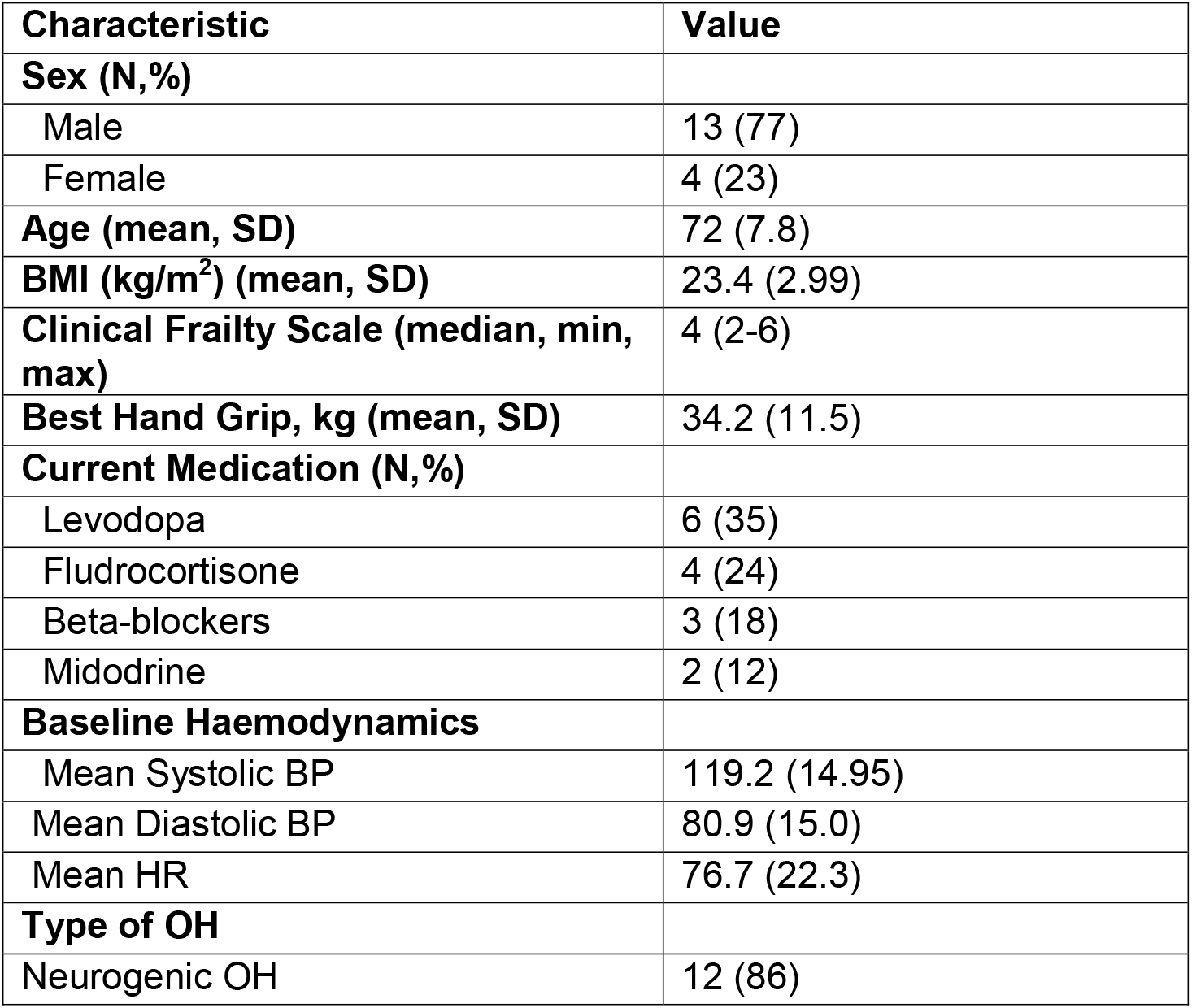
Baseline demographic, clinical and haemodynamic characteristics (N = 17) *Caption: Table 1 summarises baseline demographic, clinical and haemodynamic characteristics for the cohort (N=17) at the supine, unassisted assessment. Participants were older (mean age 72 years) and predominantly male (77%), with a mean BMI of 23*.*4 kg/m*^*2*^ *and a median Clinical Frailty Scale score of 4 (range 2–6). Mean dominant-hand grip strength was 34*.*2 kg, and most had neurogenic orthostatic hypotension (86%). Baseline supine haemodynamics were: systolic blood pressure 119*.*2 mmHg, diastolic blood pressure 80*.*9 mmHg and heart rate 76*.*7 bpm. Concomitant medications included levodopa (35%), fludrocortisone (24%), beta-blockers (18%) and midodrine (12%). Continuous data are mean (SD) unless stated; medication categories are not mutually exclusive*.

**TABLE 2.** Primary outcome. Responder rate and blood pressure change versus unassisted active stand, by timing of device activation (N=14) *Caption: Primary outcome for the fixed-sequence study. Columns show device activation while supine before standing (pre-stand) and immediately after standing (post-stand). A responder was defined a priori as achieving, with device use versus the unassisted active stand*, ≥*10 mmHg improvement in any one of: resting blood pressure; nadir standing blood pressure at 30– 60 s; or reduction in the orthostatic blood-pressure drop (systolic or diastolic). Proportions are tested against a 20% benchmark using an exact binomial test; 95% CIs are Wilson. Blood-pressure changes are within-participant differences among responders, derived from continuous signals processed with a 10-s moving average*.

| Characteristic | Device activated while supine (pre-stand) | Device activated immediately after standing (post-stand) |
| --- | --- | --- |
| Responders, n/N (%), 95% CI | 3/14 (21%); 8–48% | 6/14 (43%); 21–67% |
| Exact binomial test vs 20% benchmark | $p = 0.30$ | $p = 0.012$ |
| $\Delta$ Systolic BP, mmHg (mean [SD]) among responders | 18.58 (7.05) | 22.13 (16.29) |
| $\Delta$ Diastolic BP, mmHg (mean [SD]) among responders | 14.02 (11.51) | 19.52 (12.47) |

- Analyses use a 10-second moving average for continuous BP. - Benchmark is 20% responder rate based on standard non-pharmacological care. - 95% CIs shown are Wilson; exact binomial p values test proportion >20%. - BP changes are within-person differences vs the unassisted condition, calculated among responders only.

