## Supplementary Figure 1 for "ISO-101, a Wrist-Worn Isometric Counter-Manoeuvre Device, Improves Blood Pressure Recovery in Orthostatic Hypotension"

### Slide 1
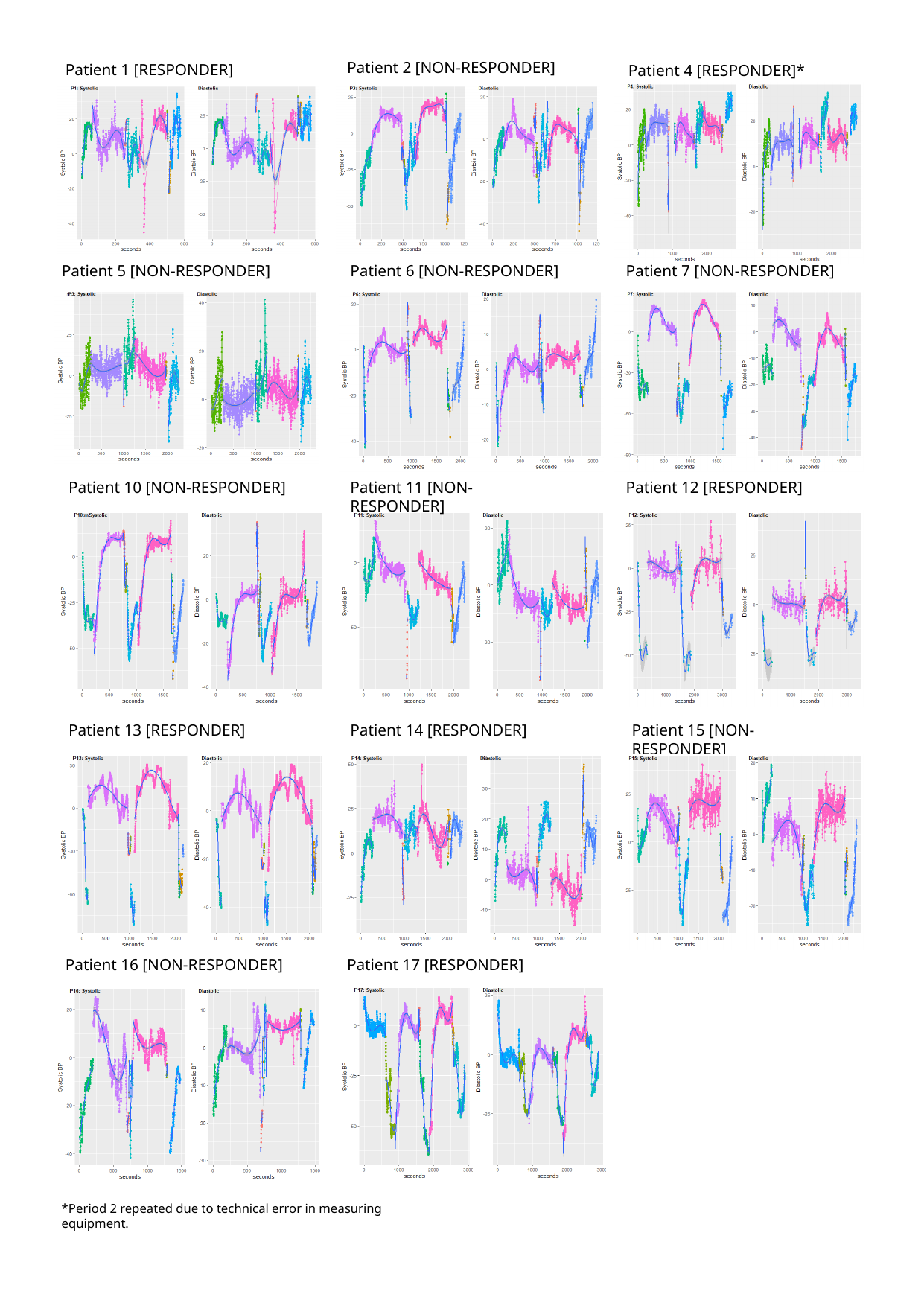

Patient 2 [NON-RESPONDER]
Patient 1 [RESPONDER]
Patient 4 [RESPONDER]*
Patient 5 [NON-RESPONDER]
Patient 6 [NON-RESPONDER]
Patient 7 [NON-RESPONDER]
Patient 12 [RESPONDER]
Patient 11 [NON-RESPONDER]
Patient 10 [NON-RESPONDER]
Patient 13 [RESPONDER]
Patient 15 [NON-RESPONDER]
Patient 14 [RESPONDER]
Patient 16 [NON-RESPONDER]
Patient 17 [RESPONDER]
*Period 2 repeated due to technical error in measuring equipment.
